# Negative Clinical Outcomes Between Silver Diamine Fluoride, Sedation, and General Anesthesia Treatment for Children with Early Childhood Caries: A Cohort Study

**DOI:** 10.1101/2023.12.15.23300046

**Authors:** David Okuji, Yen Dinh, Tony McClure, Myeonggyun Lee

**Affiliations:** Senior Associate Director, NYU Langone Dental Medicine, Hansjörg Wyss Department of Plastic Surgery, Division of Dental Medicine, NYU Grossman School of Medicine, 5800 Third Avenue, 3^rd^ Floor, Brooklyn, NY 11220; Predoctoral Student, Harvard School of Dental Medicine, 188 Longwood Avenue, Boston, MA 02115; Associate Director, NYU Langone Dental Medicine, Southcentral Foundation/Alaska Native Medical Center, 4441 Diplomacy Drive, Anchorage, Alaska 99508; Biostatistician, Division of Biostatistics, Department of Population Health, NYU Grossman School of Medicine, 180 Madison Ave., 5th Floor, New York, NY 10016

**Keywords:** early-childhood-caries, silver-diamine-fluoride, sedation, general-anesthesia, outcomes

## Abstract

**Purpose:** This study compared the wait-time for treatment completion and pre- and post-treatment outcomes of treating early childhood caries with silver diamine fluoride, sedation, and general anesthesia.

**Methods:** This retrospective study examined children with early childhood caries treated with either silver diamine fluoride, sedation, or general anesthesia at federally qualified health centers. Demographics, wait-time for treatment completion, and pre- and post-treatment clinical outcomes were compared with analysis of variance for continuous variables and the Chi-square test for categorical variables. This study was reviewed and approved by Southcentral Foundation Research Review.

**Results:** The outcomes between the silver diamine fluoride, sedation, and general anesthesia groups were respectively: 1) average wait-times to complete treatment at 49.6, 62.5, and 116.3 days, 2) mean number of pre-treatment visits at 1.08, 1.25, and 1.61, 3) mean number of post-treatment visits at 1.41, 1.29, and 1.45.

Multiple negative outcomes were identified when the sedation and general anesthesia groups were compared with the silver diamine fluoride reference group for 1) pre-treatment visits with un-planned visits (for general anesthesia only), pain, intra-oral swelling, and prescriptions for pain and antibiotic medications (general anesthesia only) and 2) post-treatment visits with new carious lesions on permanent molars, new carious lesions on primary teeth (sedation only), intra-oral swelling (sedation only only), broken restorations, displaced restorations, and pulpal therapy (sedation only).

**Conclusions:** Silver diamine fluoride provides timely and effective caries management with lower wait-time for treatment completion, clinical outcomes consistent with minimally invasive treatment, and mitigation for the risk of negative pre- and post-operative clinical outcomes compared to treatment under sedation or general anesthesia.

## Introduction

According to the 2016 Morbidity and Mortality Weekly Report from the Centers for Disease Control and Prevention, caries is one of the most prevalent chronic diseases among children in the United States.^1^ Early childhood caries (**ECC**) refers to carious lesions in infants and preschool children. ECC has severe negative impacts on young children. ECC causes pain, infection, difficulty eating, poor weight gain, and behavioral issues when left untreated.^2^ The American Academy of Pediatric Dentistry defines ECC as one or more decayed, missing, or filled tooth surfaces due to caries in any primary tooth of a child under 6 years old. In children under three years old, severe early childhood caries (**S-ECC**) refers to any signs of tooth decay on smooth tooth surfaces. For children age 3-5, S-ECC is when there is one or more cavitated, missing, or filled smooth surfaces in primary maxillary anterior teeth or decayed, missing, or filled score ≥4 (age 3), ≥5 (age 4), or ≥6 (age 5).^3^

Dentists treat ECC and S-ECC through a combination of preventive measures and restorative treatments. Despite numerous effective treatment options, however, challenges often arise because young patients have fear of the dentist, or are unable to understand and cooperate for the duration of the procedure.^4^ For these patients, pharmacological management ranging from minimal sedation (**SED**) to general anesthesia (**GA**) has been the standard of care to facilitate a safer environment for patients to receive comprehensive, high-quality dental care.^2^

While these time-efficient methods reduce patient discomfort and enable clinicians to provide complete dental treatment, they also come with risks that require careful consideration. Sedation and GA in young children can result in allergic reactions, respiratory complications, cardiovascular issues, and in some cases, even death.^5–9^ Studies on animal models have shown that when exposed to sedation and GA for prolonged periods or repeatedly during early development, neurons may undergo apoptosis, resulting in persistent functional impairments that affect learning, memory, and behavior.^10^ According to clinical studies, repeated exposure to GA before the age of two has been identified as a significant risk factor for the development of learning disorders and attention-deficit/hyperactivity disorder.^11,12^

While sedation and GA provide an optimal environment for dental treatment, they can have high failure rates for restoration in young children with ECC. Studies indicate that parents do not recognize the importance of regular dental check-ups after sedation or GA procedures, resulting in a lack of follow-up care.^13^ Factors such as nursing bottle time after GA, duration and techniques children use to brush their teeth, and inadequate dental care contribute to the need for repeat treatment under sedation or GA.^14^ Furthermore, children with ECC are particularly vulnerable to the development of new and recurrent carious lesions. Lin et al. conducted a cross-sectional study involving 79 pediatric patients who previously received treatment under GA for ECC, which found that 79.7 percent of the children had developed new carious lesions at the 12-month follow-up examination.^15^

Access to sedation and GA services can be challenging for some children due to long wait times, limited insurance coverage, high demand for GA services, reduced operating room capacity, and a shortage of trained clinicians to perform GA. Studies have shown that longer wait times for dental procedures are associated with a number of negative outcomes, including increased pain, a higher number of teeth treated than originally planned, and more frequent pre- and post-operative visits.^16^ Another significant barrier occurs when insurance companies refuse to reimburse for sedation and GA dental procedure.^17^ Patients are often left to bear the burden of these out-of-pocket costs, which can range from hundreds to thousands of dollars, disproportionately affecting low-income patients who already face significant barriers to accessing dental care.^18^

Due to the aforementioned drawbacks of sedation and GA, alternative approaches, such as minimally invasive care with silver diamine fluoride (**SDF**), have gained increasing attention in recent years for their potential to provide non-invasive, effective treatment with fewer associated risks and costs. SDF is the most concentrated fluoride product commercially available for caries management, containing approximately 25 percent silver, eight percent amine, five percent fluoride, and 62 percent water.^19^ Multiple clinical trials and systematic reviews have demonstrated the effectiveness of SDF in arresting or halting the progression of carious lesions while promoting remineralization of the surface zone.^20–23^

SDF is applied topically with a micro-brush. The resulting arrested lesion is stained black, with no other major complications reported.^21^ Although staining, particularly on anterior teeth, is undesirable, most parents still prefer SDF over the use of sedation and general anesthesia.^24^ SDF has also demonstrated greater efficacy in preventing new carious lesions compared to topical fluoride, while being a more cost-effective alternative to sealants.^25^ As its potential benefits appear to outweigh the drawbacks, researchers and clinicians are increasingly advocating for the use of SDF.^26–28^ However, additional research on the clinical outcomes of patients is necessary to gain a comprehensive understanding of the effectiveness and potential long-term effects of SDF compared to the current standard of care.

This present study aims to compare the wait-time for treatment completion and negative clinical outcomes of treating children with ECC with SDF, sedation, or GA treatment modalities; with the null hypothesis that there are no differences in pre-treatment and post-treatment negative clinical outcomes.

## Methods

### Study Design & Setting

A cohort study design with a retrospective chart review was conducted for children with ECC who were treated with either SDF, under sedation, or under GA at NYU Langone Dental Medicine-Advanced Education in Pediatric Dentistry program training locations in seven states (Alaska, Arizona, Florida, Hawaii, Massachusetts, Missouri, New York, and Tennessee) between January 1, 2010, and April 24, 2022. The study was approved by the NYU Grossman School of Medicine Institutional Review Board under protocol number 20-00142.

### Participants, Variables, Data Sources/Measurement

Eligible participants were included as follows: 1) patients of record at an NYU Langone Dental Program training location; 2) less than 7-years old on the date of the treatment plan for SDF, sedation, or GA; 3) American Society of Anesthesiologists (**ASA**) classification I, II, and III; 4) recorded with <4 pre-treatment visits between the treatment planning date and the treatment date; 5) SDF participants had <4 total SDF visits; 6) sedation participants had <4 total sedation visits; and 7) all participants had <6 post-treatment visits between the final treatment date and the next regularly scheduled 6-month recall visit.

Participants were assigned to their categorical variable of treatment modalities by filtering of electronic health records with Current Dental Terminology codes definitions, as follows: 1) SDF treatment as “D1354 (interim caries arresting medication),” sedation treatment as “D9248 (non-intravenous conscious sedation),” and general anesthesia treatment as “D9420 (Hospital Call).”

For cases where invasive procedures are not feasible or suitable, SDF is a minimally invasive care treatment employed to manage and halt the progression of dental caries during the patient’s visit. For patients with anxiety and fear during the dental procedure, various levels of SED are utilized to induce a relaxed and calm state while keeping the patient conscious and responsive throughout the treatment. GA induces a state of unconsciousness and is primarily employed for complicated or invasive procedures, as well as for patients with special health care needs or complex medical conditions, extreme anxiety and fear, or limited cooperation.

Data obtained from each chart included demographic information such as age, sex, ethnicity, race, payer source, date of initial treatment plan consultation, date of treatment plan completion, and pre- and post-treatment data. The treatment data included ASA classification; purpose of visits; new carious lesions on primary and permanent teeth; pain, intra-oral and extra-oral swelling; analgesic and antibiotic prescriptions; extractions; intra-oral incision and drainage; pulpal therapy; definitive, broken, and displaced restorations; SDF topical application; and interim therapeutic restorations.

### Bias

Information bias, due to missing chart information, was addressed by excluding incomplete records from the analysis under the missing at random assumption since there was a sufficiently large number of participant observations. Confounding bias was addressed with adjustments in the statistical analyses for the confounding variables of child’s age, gender, race/ethnicity, and payor source.

### Study Size

A power analysis based upon the results for the effect size for appointment wait-times from a similar study,^16^ estimated a sample size of 805 participants to yield an 80 percent power level at alpha equal to 0.05. The sample size for this present study exceeded that of the power analysis estimate to ensure a sufficient statistical power level.

### Quantitative Variables

Wait-time for treatment completion (in days) was defined as a continuous variable from the date of the initial treatment plan consultation to the date of completed intervention.

The negative pre- and post-treatment clinical outcomes were defined “Yes” responses as follows:

1. New carious lesions on permanent molars
2. New carious lesions on primary teeth
3. Presence of pain
4. Presence of extra-oral swelling
5. Presence of intra-oral swelling
6. Presence of broken restorations and/or space maintainers (i.e., “broken” restorations/space maintainers included those with marginal breakdown, deep marginal ditching, tooth discoloration, chipped or fractured restoration, tooth fracture, and secondary caries^29^)
7. Observation of displaced restoration(s) and/or space maintainers (i.e., partial- or full-displacement)
8. Prescription for pain medication(s)
9. Prescription for antibiotic medication(s)
10. Performance of dental extraction(s)
11. Performance of incision and drainage (intra-oral only)
12. Performance of pulpal therapy
13. Performance of definitive restoration(s)
14. Performance of SDF topical application
15. Performance of interim therapeutic restoration(s)

In addition, the negative post-treatment clinical outcomes included the additional variable as follows:

Purpose of visit category checked as “Planned,” “Unplanned,” or “Other.”

### Statistical Methods

For descriptive statistics, mean and standard deviation for continuous variables and frequency and percentage for categorical variables were calculated. The descriptive statistics were reported overall in the pooled data. Each variable, including demographics, wait-time for treatment completion, and clinical outcomes, were compared across the three treatment groups with the analysis of variance for continuous variables and the Chi-square test for categorical variables. Because the variable for wait-time for treatment completion was right-skewed, the reported median with interquartile range and the p-value using Kruskal-Wallis test as a nonparametric test were additionally reported.

To compare wait-time for treatment completion between the three groups, adjusted for potential confounders, a generalized linear model with the log of mean response (i.e., Gamma model with the log link) where the waiting time outcome is right-skewed was conducted. Logistic regressions for each question from pre- and post-treatment visits were conducted to identify the association of treatment while adjusting for the confounders.

All statistical tests were two-sided, with Wald type confidence intervals provided, and p<0.05 was considered statistically significant. All analyses were performed in R version 4.2.1.^30^

## Results

Table 1 presents the descriptive analyses and statistically significant inferential associations across three treatment modalities. Descriptively, the sample size was 4,730 child-participants, with a mean age of 3.9 years old, 52.5 percent of male, 46.0 percent Hispanic, 13.6 percent White, and 90.5 percent with Medicaid insurance. The GA group had the highest number of pre-treatment visits (mean of 1.61), followed by the SED (1.25) and SDF (1.08) groups with p<0.001.

**Table 1:** Demographic and main effect outcomes between SDF, SED, and GA treatment modalities.

|  | Overall | SDF | SED | GA | p-value |
| --- | --- | --- | --- | --- | --- |
| n (%) | 4730 (100.0) | 1171 (24.8) | 1642 (34.7) | 1917 (40.5) |  |
| Child's age in years; mean (sd); Continuous | 3.98 (1.26) | 3.43 (1.30) | 4.34 (1.18) | 3.99 (1.18) | <0.001 |
| Sex (%); Nominal |  |  |  |  | 0.131 |
| Male | 2483 (52.5) | 623 (53.2) | 827 (50.4) | 1033 (53.9) |  |
| Female | 2238 (47.3) | 547 (46.7) | 809 (49.3) | 882 (46.0) |  |
| Other | 3 ( 0.1) | 1 ( 0.1) | 2 ( 0.1) | 0 ( 0.0) |  |
| No response | 6 ( 0.1) | 0 ( 0.0) | 4 ( 0.2) | 2 ( 0.1) |  |
| Child's Race/Ethnicity n (%); Nominal |  |  |  |  | <0.001 |
| Hispanic | 2174 (46.0) | 674 (57.6) | 687 (41.8) | 813 (42.4) |  |
| White, Non-Hispanic | 641 (13.6) | 141 (12.0) | 193 (11.8) | 307 (16.0) |  |
| Black or African American, Non-Hispanic | 275 ( 5.8) | 62 ( 5.3) | 101 ( 6.2) | 112 ( 5.8) |  |
| American Indian or Alaska Native, Non-Hispanic | 183 ( 3.9) | 26 ( 2.2) | 78 ( 4.8) | 79 ( 4.1) |  |
| Asian, Non-Hispanic | 170 ( 3.6) | 47 ( 4.0) | 51 ( 3.1) | 72 ( 3.8) |  |
| Native Hawaiian or Other Pacific Islander, Non-Hispanic | 353 ( 7.5) | 89 ( 7.6) | 160 ( 9.7) | 104 ( 5.4) |  |
| Two or more races, Non-Hispanic | 108 ( 2.3) | 41 ( 3.5) | 29 ( 1.8) | 38 ( 2.0) |  |
| No response | 826 (17.5) | 91 ( 7.8) | 343 (20.9) | 392 (20.4) |  |
| ASA Classification; n (%); Nominal |  |  |  |  | <0.001 |
| ASA 1 | 4048 (85.6) | 1021 (87.2) | 1440 (87.7) | 1587 (82.8) |  |
| ASA 2 | 636 (13.4) | 142 (12.1) | 200 (12.2) | 294 (15.3) |  |
| ASA 3 | 46 ( 1.0) | 8 ( 0.7) | 2 ( 0.1) | 36 ( 1.9) |  |
| Payor Source; n (%); Nominal |  |  |  |  | <0.001 |
| Medicaid | 4279 (90.5) | 1056 (90.2) | 1455 (88.6) | 1768 (92.2) |  |
| Children's Health Insurance Program (CHIP) | 11 ( 0.2) | 1 ( 0.1) | 6 ( 0.4) | 4 ( 0.2) |  |
| Commercial Insurance | 208 ( 4.4) | 59 ( 5.0) | 103 ( 6.3) | 46 ( 2.4) |  |
| No Insurance | 78 ( 1.6) | 36 ( 3.1) | 20 ( 1.2) | 22 ( 1.1) |  |
| Multiple insurance coverage | 31 ( 0.7) | 7 ( 0.6) | 11 ( 0.7) | 13 ( 0.7) |  |
| No response | 123 ( 2.6) | 12 ( 1.0) | 47 ( 2.9) | 64 ( 3.3) |  |
| Average number of pre-treatment visits; mean (SD) | 1.41 (0.78) | 1.08 (0.34) | 1.25 (0.62) | 1.61 (0.90) | <0.001 |
| Average number of post-treatment visits; mean (SD) | 1.39 (0.73) | 1.41 (0.83) | 1.29 (0.63) | 1.45 (0.73) | <0.001 |
| waiting time, mean (SD) | 81.07<br>(100.63) | 49.57<br>(51.27) | 62.45<br>(88.62) | 116.27<br>(120.50) | <0.001 |
| waiting time, median (IQR) | 49 (72) | 38 (39) | 42 (55) | 77 (116) | <0.001* |

In post-treatment visits, the GA group had the highest number (1.45), followed by SDF (1.41) and SED (1.29) with p<0.001. The hierarchy of mean average wait-times for treatment completion, in days, was GA (116.3), SED (62.5), and SDF (49.6) with p<0.001. After adjusting for potential confounders, the wait-times for treatment completion for the GA and SED groups were respectively 2.36 (= exp (0.86)) and 1.30 (= exp (0.26)) times higher compared to SDF group with p<0.001 (Figure 1).

**Figure 1:**
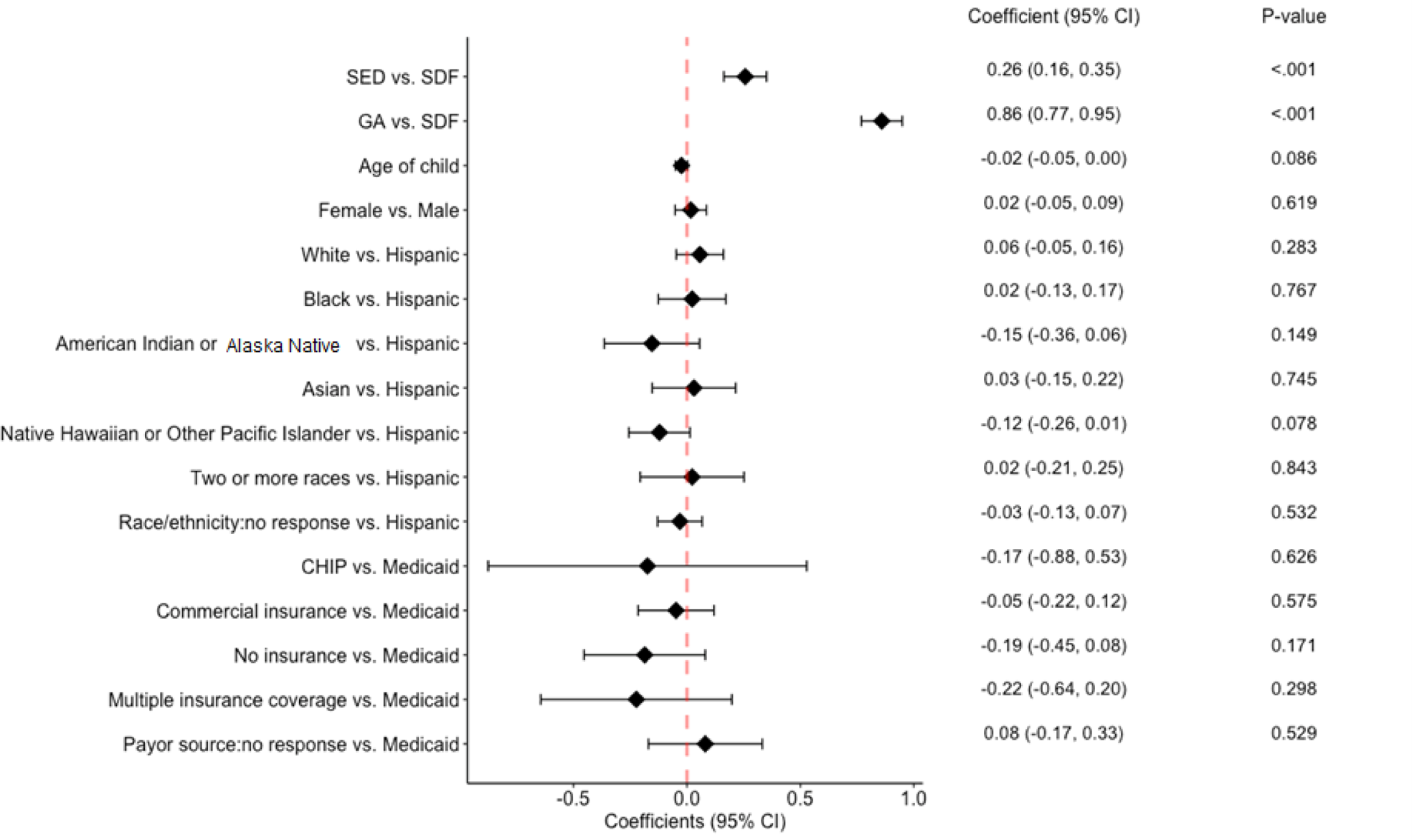
Linear regression of wait-time for treatment completion.

Table 2 provides a comprehensive breakdown of pre-treatment visits and clinical outcomes based on the three treatment modalities. The GA group had the highest proportion of unplanned visits, pain, prescription for antibiotic medications, and other procedures performed followed by SED and SDF (p<0.001). Additionally, the GA group also had the highest proportion of prescriptions for pain medication, dental extraction(s), and interim therapeutic restoration(s) followed by SDF and SED, respectively (p<0.001). The SED group had the highest proportion of at least one erupted permanent molar and intra-oral swelling followed by GA and SDF (p<0.001). The SDF group had the highest proportion of new caries on primary teeth and exam, prophy, topical fluoride, and/or radiograph procedures followed by GA and SED (p<0.001). Furthermore, the SDF also had the highest proportion of pulpal therapy and definitive restoration procedures followed by SED and GA (p<0.001). In terms of intra-oral incision and drainage procedures, SED and GA had the highest proportion, followed by SDF (p<0.001).

**Table 2:** Pre-treatment clinical outcomes between SDF, SED, and GA treatment modalities.

| **PRE-TREATMENT VISITS** | **Overall** | **SDF** | **SED** | **GA** | **p-value** |
| --- | --- | --- | --- | --- | --- |
| **N** | 2141 | 293 | 541 | 1307 |  |
| Purpose of pre-treatment visit; Nominal; n (%) |  |  |  |  | <0.001 |
| Planned visit | 1854 (86.6) | 270 (92.2) | 495 (91.5) | 1089 (83.3) |  |
| Unplanned visit | 265 (12.4) | 12 ( 4.1) | 42 ( 7.8) | 211 (16.1) |  |
| Other | 4 ( 0.2) | 1 ( 0.3) | 0 ( 0.0) | 3 ( 0.2) |  |
| No Response | 18 ( 0.8) | 10 ( 3.4) | 4 ( 0.7) | 4 ( 0.3) |  |
| Is at least one permanent molar partially or fully erupted? Nominal; n (%) |  |  |  |  | <0.001 |
| No | 1838 (85.8) | 262 (89.4) | 430 (79.5) | 1146 (87.7) |  |
| Yes | 214 (10.0) | 18 ( 6.1) | 88 (16.3) | 108 ( 8.3) |  |
| No Response | 89 ( 4.2) | 13 ( 4.4) | 23 ( 4.3) | 53 ( 4.1) |  |
| New carious lesions on the partially or fully erupted permanent molar? Nominal; n (%) |  |  |  |  | 0.364 |
| No | 1962 (91.6) | 267 (91.1) | 501 (92.6) | 1194 (91.4) |  |
| Yes | 25 ( 1.2) | 1 ( 0.3) | 8 ( 1.5) | 16 ( 1.2) |  |
| No Response | 154 ( 7.2) | 25 ( 8.5) | 32 ( 5.9) | 97 ( 7.4) |  |
| New carious lesion on primary tooth/teeth? Nominal; n (%) |  |  |  |  | <0.001 |
| No | 1709 (79.8) | 119 (40.6) | 478 (88.4) | 1112 (85.1) |  |
| Yes | 359 (16.8) | 164 (56.0) | 48 ( 8.9) | 147 (11.2) |  |
| No Response | 73 ( 3.4) | 10 ( 3.4) | 15 ( 2.8) | 48 ( 3.7) |  |
| Presence of pain? Nominal; n (%) |  |  |  |  | <0.001 |
| No | 1666 (77.8) | 270 (92.2) | 414 (76.5) | 982 (75.1) |  |
| Yes | 432 (20.2) | 13 ( 4.4) | 117 (21.6) | 302 (23.1) |  |
| No Response | 43 ( 2.0) | 10 ( 3.4) | 10 ( 1.8) | 23 ( 1.8) |  |
| Presence of Extraoral swelling? Nominal; n (%) |  |  |  |  | 0.167 |
| No | 2057 (96.1) | 280 (95.6) | 524 (96.9) | 1253 (95.9) |  |
| Yes | 36 ( 1.7) | 3 ( 1.0) | 5 ( 0.9) | 28 ( 2.1) |  |
| No Response | 48 ( 2.2) | 10 ( 3.4) | 12 ( 2.2) | 26 ( 2.0) |  |
| Presence of Intraoral swelling? Nominal; n (%) |  |  |  |  | <0.001 |
| No | 1878 (87.7) | 273 (93.2) | 455 (84.1) | 1150 (88.0) |  |
| Yes | 213 ( 9.9) | 10 ( 3.4) | 74 (13.7) | 129 ( 9.9) |  |
| No Response | 50 ( 2.3) | 10 ( 3.4) | 12 ( 2.2) | 28 ( 2.1) |  |
| Presence of broken restorations(s) and/or space maintainer(s)?<br>Nominal; n (%) |  |  |  |  | 0.47 |
| No | 2052 (95.8) | 283 (96.6) | 518 (95.7) | 1251 (95.7) |  |
| Yes | 8 ( 0.4) | 0 ( 0.0) | 4 ( 0.7) | 4 ( 0.3) |  |
| No Response | 81 ( 3.8) | 10 ( 3.4) | 19 ( 3.5) | 52 ( 4.0) |  |
| Observation of displaced restoration(s) and/or space maintainer(s)?<br>Nominal; n (%) |  |  |  |  | 0.275 |
| No | 2051 (95.8) | 283 (96.6) | 517 (95.6) | 1251 (95.7) |  |
| Yes | 12 ( 0.6) | 0 ( 0.0) | 6 ( 1.1) | 6 ( 0.5) |  |
| No Response | 78 ( 3.6) | 10 ( 3.4) | 18 ( 3.3) | 50 ( 3.8) |  |
| Prescription for pain medication(s)? Nominal; n (%) |  |  |  |  | <0.001 |
| No | 2078 (97.1) | 282 (96.2) | 535 (98.9) | 1261 (96.5) |  |
| Yes | 44 ( 2.1) | 1 ( 0.3) | 1 ( 0.2) | 42 ( 3.2) |  |
| No Response | 19 ( 0.9) | 10 ( 3.4) | 5 ( 0.9) | 4 ( 0.3) |  |
| Prescription for antibiotic medication(s)? Nominal; n (%) |  |  |  |  | <0.001 |
| No | 2022 (94.4) | 277 (94.5) | 520 (96.1) | 1225 (93.7) |  |
| Yes | 99 ( 4.6) | 6 ( 2.0) | 15 ( 2.8) | 78 ( 6.0) |  |
| No Response | 20 ( 0.9) | 10 ( 3.4) | 6 ( 1.1) | 4 ( 0.3) |  |
| Dental extraction(s) performed? Nominal; n (%) |  |  |  |  | <0.001 |
| No | 2011 (93.9) | 270 (92.2) | 517 (95.6) | 1224 (93.6) |  |
| Yes | 111 ( 5.2) | 13 ( 4.4) | 19 ( 3.5) | 79 ( 6.0) |  |
| No Response | 19 ( 0.9) | 10 ( 3.4) | 5 ( 0.9) | 4 ( 0.3) |  |
| Incision and drainage (intraoral only) performed? Nominal; n (%) |  |  |  |  | <0.001 |
| No | 2119 (99.0) | 283 (96.6) | 535 (98.9) | 1301 (99.5) |  |
| Yes | 3 ( 0.1) | 0 ( 0.0) | 1 ( 0.2) | 2 ( 0.2) |  |
| No Response | 19 ( 0.9) | 10 ( 3.4) | 5 ( 0.9) | 4 ( 0.3) |  |
| Pulpal therapy performed? Nominal; n (%) |  |  |  |  | <0.001 |
| No | 2106 (98.4) | 277 (94.5) | 533 (98.5) | 1296 (99.2) |  |
| Yes | 16 ( 0.7) | 6 ( 2.0) | 3 ( 0.6) | 7 ( 0.5) |  |
| No Response | 19 ( 0.9) | 10 ( 3.4) | 5 ( 0.9) | 4 ( 0.3) |  |
| Definitive restoration(s) performed? Nominal; n (%) |  |  |  |  | <0.001 |
| No | 2042 (95.4) | 250 (85.3) | 516 (95.4) | 1276 (97.6) |  |
| Yes | 81 ( 3.8) | 33 (11.3) | 21 ( 3.9) | 27 ( 2.1) |  |
| No Response | 18 ( 0.8) | 10 ( 3.4) | 4 ( 0.7) | 4 ( 0.3) |  |
| Interim Therapeutic Restoration(s)-ITR performed? Nominal; n (%) |  |  |  |  | <0.001 |
| No | 2072 (96.8) | 277 (94.5) | 528 (97.6) | 1267 (96.9) |  |
| Yes | 51 ( 2.4) | 6 ( 2.0) | 9 ( 1.7) | 36 ( 2.8) |  |
| No Response | 18 ( 0.8) | 10 ( 3.4) | 4 ( 0.7) | 4 ( 0.3) |  |
| Exam, prophylaxis, topical fluoride, and/or radiographs performed? Nominal; n (%) |  |  |  |  | <0.001 |
| No | 982 (45.9) | 106 (36.2) | 293 (54.2) | 583 (44.6) |  |
| Yes | 1141 (53.3) | 177 (60.4) | 244 (45.1) | 720 (55.1) |  |
| No Response | 18 ( 0.8) | 10 ( 3.4) | 4 ( 0.7) | 4 ( 0.3) |  |
| Other procedure(s) performed? Nominal; n (%) |  |  |  |  | <0.001 |
| No | 1665 (77.8) | 262 (89.4) | 427 (78.9) | 976 (74.7) |  |
| Yes | 457 (21.3) | 21 ( 7.2) | 109 (20.1) | 327 (25.0) |  |
| No Response | 19 ( 0.9) | 10 ( 3.4) | 5 ( 0.9) | 4 ( 0.3) |  |

Table 3 outlines the post-treatment visits and the corresponding clinical outcomes for each of the three treatment modalities. The SED group had the highest proportion of exam, prophy, topical fluoride and/or radiographs performed, as well as at least one erupted permanent molar and new carious lesion on permanent tooth, followed by GA and SDF (p<0.001). The SED group also had the highest proportion of new caries on primary teeth, prescription for antibiotic medication, presence of pain, intra-oral swellings, and pulpal therapy performed followed by SDF and GA, respectively (p<0.05). The GA group had the highest proportion of broken or displaced restoration(s)/space maintainer(s) followed by SED and SDF (p<0.001). The GA group also had the highest proportion of “other” procedures performed followed by SDF and SED (p<0.001). In terms of prescriptions for pain medication, GA and SDF had the highest proportion, followed by SED (p<0.001). The SDF group had the highest proportion of dental extraction(s), definitive restoration(s), SDF topical application, and interim therapeutic restoration(s) performed, followed by SED and GA (p<0.001).

**Table 3:** Post-treatment clinical outcomes between SDF, SED, and GA treatment modalities.

| **POST-TREATMENT VISITS** | **Overall** | **SDF** | **SED** | **GA** | **p-value** |
| --- | --- | --- | --- | --- | --- |
| N | 6367 | 1607 | 2070 | 2690 |  |
| Purpose of post-treatment visit; Nominal; n (%) |  |  |  |  | 0.132 |
| Planned visit | 5901 (93.1) | 1490 (94.1) | 1910 (92.4) | 2501 ( 93.0) |  |
| Unplanned visit | 431 ( 6.8) | 90 ( 5.7) | 155 ( 7.5) | 186 ( 6.9) |  |
| Other | 8 ( 0.1) | 4 ( 0.3) | 1 ( 0.0) | 3 ( 0.1) |  |
| No Response | 1 ( 0.0) | 0 ( 0.0) | 1 ( 0.0) | 0 ( 0.0) |  |
| Is at least one permanent molar partially or fully erupted?<br>Nominal; n (%) |  |  |  |  | <0.001 |
| No | 5054 (79.7) | 1412 (89.1) | 1505 (72.8) | 2137 ( 79.4) |  |
| Yes | 1182 (18.6) | 143 ( 9.0) | 526 (25.5) | 513 ( 19.1) |  |
| No Response | 104 ( 1.6) | 29 ( 1.8) | 35 ( 1.7) | 40 ( 1.5) |  |
| New carious lesions on the partially or fully erupted permanent<br>molar? Nominal; n (%) |  |  |  |  | <0.001 |
| No | 5498 (86.7) | 1229 (77.6) | 1868 (90.4) | 2401 ( 89.3) |  |
| Yes | 167 ( 2.6) | 16 ( 1.0) | 84 ( 4.1) | 67 ( 2.5) |  |
| No Response | 675 (10.6) | 339 (21.4) | 114 ( 5.5) | 222 ( 8.3) |  |
| New carious lesion on primary tooth/teeth? Nominal; n (%) |  |  |  |  | <0.001 |
| No | 4970 (78.4) | 1096 (69.2) | 1398 (67.7) | 2476 ( 92.0) |  |
| Yes | 1354 (21.4) | 486 (30.7) | 660 (31.9) | 208 ( 7.7) |  |
| No Response | 16 ( 0.3) | 2 ( 0.1) | 8 ( 0.4) | 6 ( 0.2) |  |
| Presence of pain? Nominal; n (%) |  |  |  |  | 0.005 |
| No | 5801 (91.5) | 1451 (91.6) | 1858 (89.9) | 2492 ( 92.6) |  |
| Yes | 526 ( 8.3) | 132 ( 8.3) | 200 ( 9.7) | 194 ( 7.2) |  |
| No Response | 13 ( 0.2) | 1 ( 0.1) | 8 ( 0.4) | 4 ( 0.1) |  |
| Presence of Extraoral swelling? Nominal; n (%) |  |  |  |  | 0.065 |
| No | 6298 (99.3) | 1573 (99.3) | 2047 (99.1) | 2678 ( 99.6) |  |
| Yes | 33 ( 0.5) | 11 ( 0.7) | 13 ( 0.6) | 9 ( 0.3) |  |
| No Response | 9 ( 0.1) | 0 ( 0.0) | 6 ( 0.3) | 3 ( 0.1) |  |
| Presence of Intraoral swelling? Nominal; n (%) |  |  |  |  | 0.002 |
| No | 6081 (95.9) | 1526 (96.3) | 1955 (94.6) | 2600 ( 96.7) |  |
| Yes | 252 ( 4.0) | 58 ( 3.7) | 106 ( 5.1) | 88 ( 3.3) |  |
| No Response | 7 ( 0.1) | 0 ( 0.0) | 5 ( 0.2) | 2 ( 0.1) |  |
| Presence of broken restorations(s) and/or space maintainer(s)?<br>Nominal; n (%) |  |  |  |  | <0.001 |
| No | 6064 (95.6) | 1576 (99.4) | 1923 (93.1) | 2565 ( 95.4) |  |
| Yes | 125 ( 2.0) | 6 ( 0.4) | 45 ( 2.2) | 74 ( 2.8) |  |
| No Response | 152 ( 2.4) | 3 ( 0.2) | 98 ( 4.7) | 51 ( 1.9) |  |
| Observation of displaced restoration(s) and/or space<br>maintainer(s)? Nominal; n (%) |  |  |  |  | <0.001 |
| No | 5949 (93.8) | 1573 (99.3) | 1883 (91.1) | 2493 ( 92.7) |  |
| Yes | 237 ( 3.7) | 9 ( 0.6) | 83 ( 4.0) | 145 ( 5.4) |  |
| No Response | 154 ( 2.4) | 2 ( 0.1) | 100 ( 4.8) | 52 ( 1.9) |  |
| Prescription for pain medication(s)? Nominal; n (%) |  |  |  |  | <0.001 |
| No | 6287 (99.2) | 1574 (99.4) | 2038 (98.6) | 2675 ( 99.4) |  |
| Yes | 31 ( 0.5) | 10 ( 0.6) | 6 ( 0.3) | 15 ( 0.6) |  |
| No Response | 22 ( 0.3) | 0 ( 0.0) | 22 ( 1.1) | 0 ( 0.0) |  |
| Prescription for antibiotic medication(s)? Nominal; n (%) |  |  |  |  | <0.001 |
| No | 6201 (97.8) | 1553 (98.0) | 1988 (96.2) | 2660 ( 98.9) |  |
| Yes | 115 ( 1.8) | 31 ( 2.0) | 55 ( 2.7) | 29 ( 1.1) |  |
| No Response | 24 ( 0.4) | 0 ( 0.0) | 23 ( 1.1) | 1 ( 0.0) |  |
| Dental extraction(s) performed? Nominal; n (%) |  |  |  |  | <0.001 |
| No | 6142 (96.9) | 1509 (95.3) | 1992 (96.4) | 2641 ( 98.2) |  |
| Yes | 197 ( 3.1) | 75 ( 4.7) | 73 ( 3.5) | 49 ( 1.8) |  |
| No Response | 1 ( 0.0) | 0 ( 0.0) | 1 ( 0.0) | 0 ( 0.0) |  |
| Incision and drainage (intraoral only) performed? Nominal; n (%) |  |  |  |  | 0.489 |
| No | 6335 (99.9) | 1582 (99.9) | 2064 (99.9) | 2689 (100.0) |  |
| Yes | 4 ( 0.1) | 2 ( 0.1) | 1 ( 0.0) | 1 ( 0.0) |  |
| No Response | 1 ( 0.0) | 0 ( 0.0) | 1 ( 0.0) | 0 ( 0.0) |  |
| Pulpal therapy performed? Nominal; n (%) |  |  |  |  | <0.001 |
| No | 6259 (98.7) | 1563 (98.7) | 2010 (97.3) | 2686 ( 99.9) |  |
| Yes | 80 ( 1.3) | 21 ( 1.3) | 55 ( 2.7) | 4 ( 0.1) |  |
| No Response | 1 ( 0.0) | 0 ( 0.0) | 1 ( 0.0) | 0 ( 0.0) |  |
| Definitive restoration(s) performed? Nominal; n (%) |  |  |  |  | <0.001 |
| No | 5993 (94.5) | 1442 (91.0) | 1912 (92.5) | 2639 ( 98.1) |  |
| Yes | 346 ( 5.5) | 142 ( 9.0) | 153 ( 7.4) | 51 ( 1.9) |  |
| No Response | 1 ( 0.0) | 0 ( 0.0) | 1 ( 0.0) | 0 ( 0.0) |  |
| SDF topical application performed? Nominal; n (%) |  |  |  |  | <0.001 |
| No | 5991 (94.5) | 1327 (83.8) | 1998 (96.7) | 2666 ( 99.1) |  |
| Yes | 346 ( 5.5) | 256 (16.2) | 67 ( 3.2) | 23 ( 0.9) |  |
| No response | 3 ( 0.0) | 1 ( 0.1) | 1 ( 0.0) | 1 ( 0.0) |  |
| Interim Therapeutic Restoration(s)-ITR performed? Nominal; n (%) |  |  |  |  | <0.001 |
| No | 6219 (98.1) | 1494 (94.3) | 2047 (99.1) | 2678 ( 99.6) |  |
| Yes | 120 ( 1.9) | 90 ( 5.7) | 18 ( 0.9) | 12 ( 0.4) |  |
| No Response | 1 ( 0.0) | 0 ( 0.0) | 1 ( 0.0) | 0 ( 0.0) |  |
| Exam, prophylaxis, topical fluoride, and/or radiographs performed? Nominal; n (%) |  |  |  |  | <0.001 |
| \ No | 1524 (23.6) | 446 (26.7) | 397 (19.0) | 681 (25.3) |  |
| Yes | 4843 (76.4) | 1161 (73.3) | 1673 (81.0) | 2009 ( 74.7) |  |
| No Response | 0 (0.0) | 0 (0.0) | 0 (0.0) | 0 (0.0) |  |
| Other procedure(s) performed? Nominal; n (%) |  |  |  |  | <0.001 |
| No | 5788 (90.9) | 1481 (92.0) | 1920 (92.7) | 2387 (88.7) |  |
| Yes | 579 ( 9.1) | 126 ( 8.0) | 150 ( 7.3) | 303 ( 11.3) |  |
| No Response | 0 (0.0) | 0 (0.0) | 0 (0.0) | 0 (0.0) |  |

During pre-treatment visits, GA group was more likely to have an unplanned visit compared to the SDF group (Figure 2). The odds ratio (**OR**) of unplanned visits for GA vs. SDF groups was 4.22 (p<0.001). Both SED and GA groups were respectively less likely to have a new carious lesion on primary teeth compared to the SDF group (OR=0.07 and 0.10 for SED and GA groups, respectively, at p<0.001). For presence of pain and intraoral swelling, and other procedures, the SED and GA groups were more likely to answer “Yes” compared to the SDF group (Figure 2). The GA group was more likely to be “Yes” for pulpal therapy and definitive restoration than the SED and SDF groups. The GA group also had a higher proportion in prescriptions for pain medication and antibiotic medication than the SDF group, but there was no difference between the SED and SDF groups on these questions (Figure 2).

**Figure 2:**
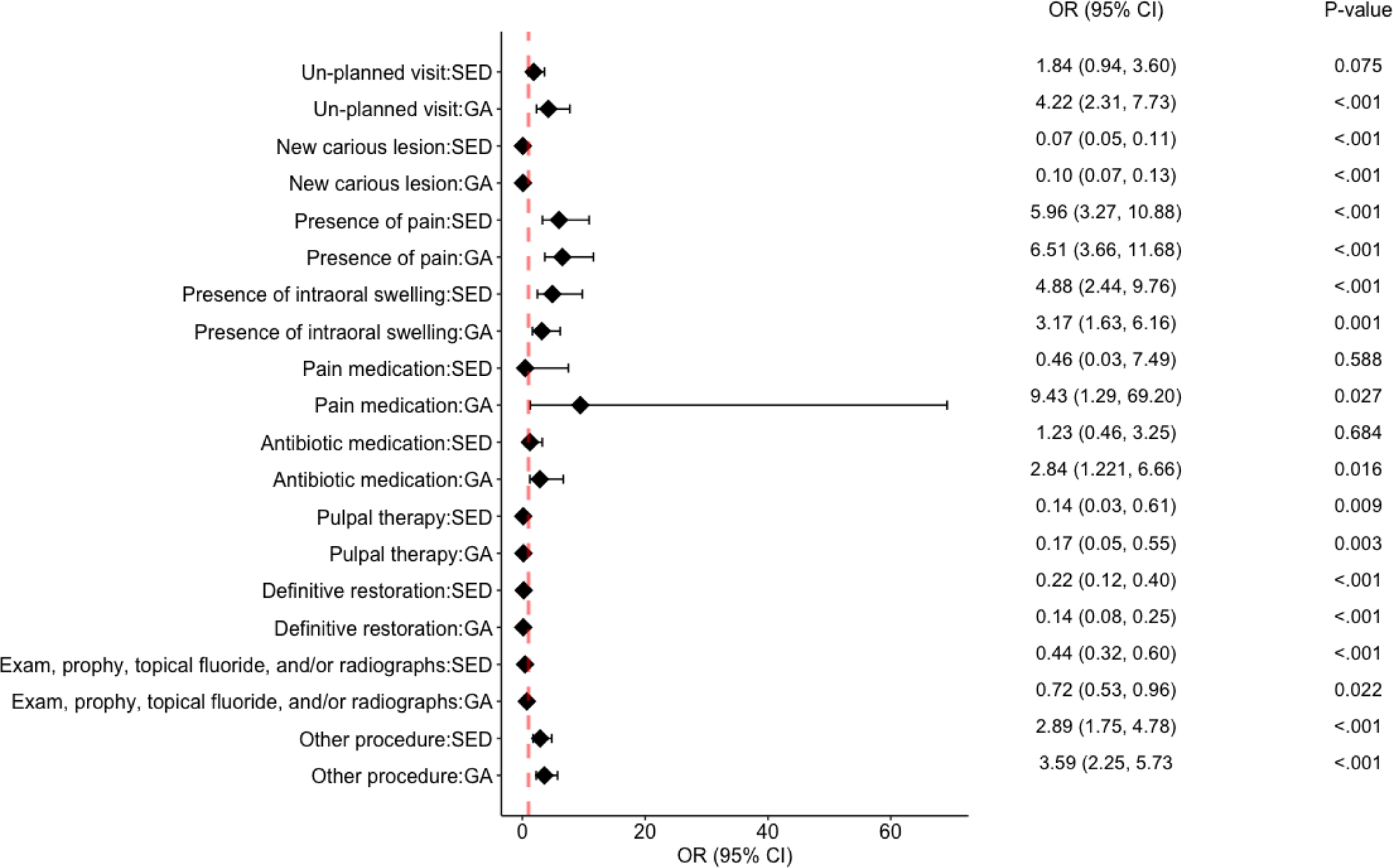
Odd ratios of pre-treatment clinical outcomes between SDF, SED, and GA treatment modalities.

For post-treatment visits, SED and GA were more likely to have an unplanned visit than the SDF group (Figure 3). Compared to the SDF group, the SED group was more likely to answer “Yes” in new carious lesions on the partially or fully erupted permanent molar, new carious lesion on primary tooth/teeth, presence of intraoral swelling and broken restoration and/or space maintainer, observation of displaced restoration and/or space maintainer, pulpal therapy, and the exam, prophy, topical fluoride, and/or radiographs questions. The GA group was less likely to answer “Yes” in definitive restoration and SDF topical application (Figure 3). For patients who had GA treatment, the presence of broken restorations and/or space maintainer, observation of displaced restoration and/or space maintainer, and other procedures were more likely to be “Yes” compared to the SDF group, while the GA group was lower in the new carious lesions on primary teeth, prescription for antibiotic medication, dental extraction, pulpal therapy, definitive restoration, and SDF topical application compared to the SDF group (Figure 3).

**Figure 3:**
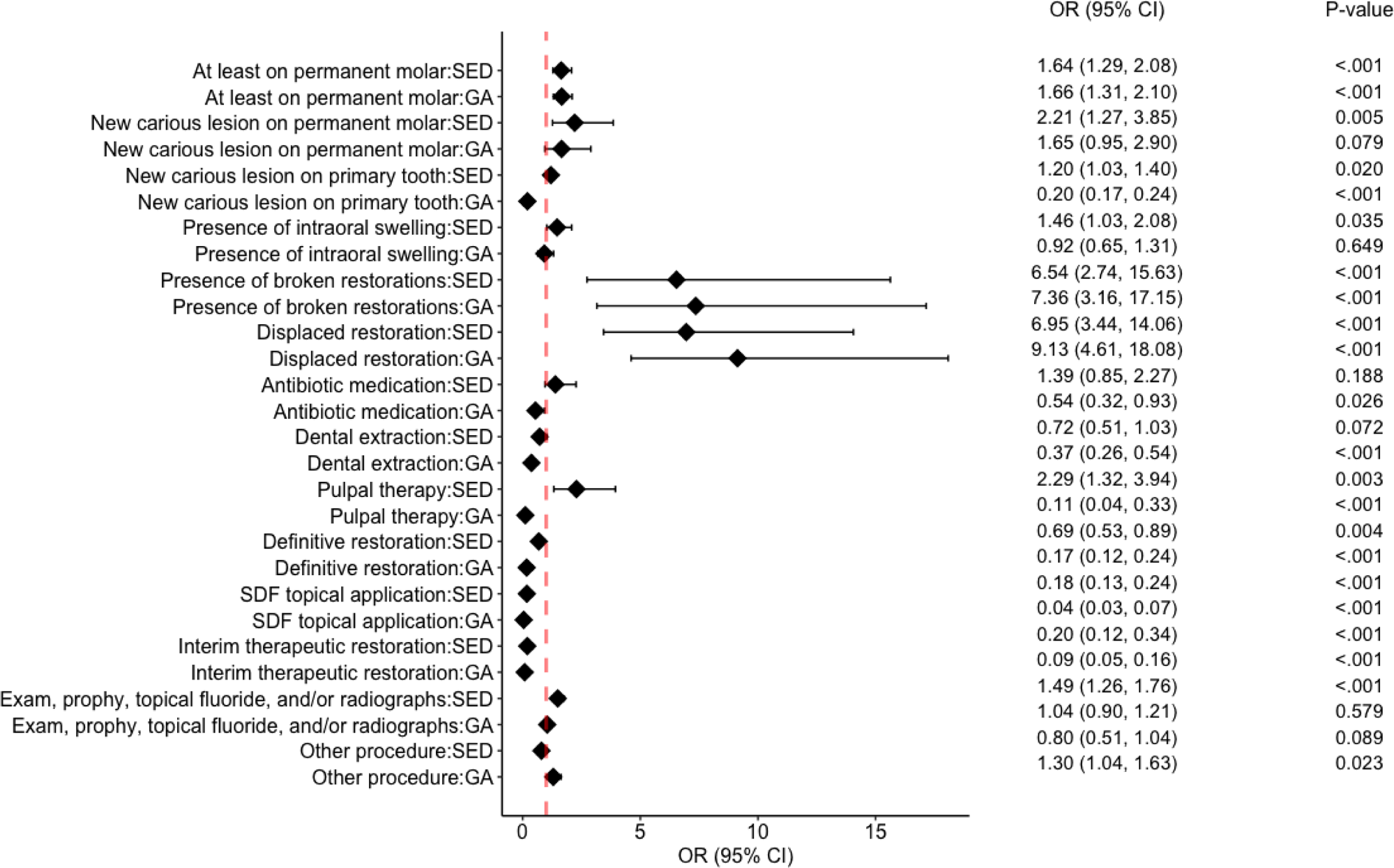
Odd ratios of post-treatment clinical outcomes between SDF, SED, and GA treatment modalities.

## Discussion

### Findings and Hypothesis, Comparison to Literature, and Significance and Implications

This study represents the first known investigation to report disparities in the clinical outcomes among SDF, SED, and GA. These disparities show that the SED and GA groups, respectively, had 1.30- and 2.36-times higher wait-times for treatment completion compared to the SDF group. As well, the SDF group, compared to the SED and GA groups, had higher odds for the clinical outcomes of 1) pre-treatment new carious lesions on primary teeth, pulpal therapy, and definitive restorations, 2) post-treatment new carious lesions on primary tooth (for GA only), intra-oral swelling (for GA only), prescription for antibiotic medication (for GA only), dental extractions (for GA only), pulpal therapy (for GA only), definitive restorations, SDF topical application, and interim therapeutic restorations. Since the primary goal of minimally invasive care is to manage the overall caries activity and stabilization of the general oral function, not the restoration of single lesions, these pre-treatment results align with the expected outcomes of the minimally invasive care approach for SDF treatment.^31^

More important, this study demonstrated multiple negative clinical outcomes when SED and GA treatments were compared with SDF treatment, such that SED and/or GA treatments have higher odds for 1) pre-treatment visits with un-planned visits (for GA only), pain, intra-oral swelling, and prescriptions for pain and antibiotic medications (for GA only) and 2) post-treatment visits with new carious lesions on permanent molars, new carious lesions on primary teeth (for SED only), intra-oral swelling (for SED only), broken restorations, displaced restorations, and pulpal therapy (for SED only) which align with the expected negative outcomes of invasive care treatment approach for SED and GA treatment due to the longer wait-times for treatment completion as reported in the existing literature.^16,32–35^ These results reject the null and accept the alternative hypothesis that there are statistically significant differences for negative clinical outcomes of SED and GA treatment when compared to SDF treatment.

Although the current literature does not include studies which directly compare SDF, SED, and GA treatment together for wait-time for treatment completion and pre- and post-treatment clinical outcomes, reported studies exist which investigate one or two of these treatment modalities separately.

The present findings are consistent with existing research demonstrating a greater wait-time for dental treatments under GA. Multiple studies have reported wait-times of 71.0, 100.1 (14.3 weeks), 110.6, 137.0, and 365.0 days.^32–33,16, 34–35^ Crystal *et al*. studied the wait-times for both sedation and general anesthesia treatments with wide ranges, respectively, from one-week to twelve-months and four-weeks to fourteen-months.^36^

With regard to pre-treatment clinical outcomes, Boehmer demonstrated determined that four- to five-year-old patients had the longest wait times for dental rehabilitation under GA, with 43 percent of them developing complications such as oral pain in the meantime.^37^ A British study found that while patients waited for treatment under GA, 41 percent had increased pain which required more analgesics and 49.4 percent required more antibiotics.^38^ This present study aligns with the findings of these studies for pre-treatment clinical outcomes.

From a post-treatment perspective, Eidelman compared clinical outcomes between SED and GA treatment and found that 74 percent of children treated under sedation required additional post-treatment compared to 59 percent of children treated under general anesthesia, with the main reasons for additional treatment due to new carious lesions, defective restorations (i.e., marginal adaptation and anatomic form), and secondary caries.^39^ Berkowitz reported new carious lesions in over 50 percent of children six months after GA treatment.^40^ For GA treatment, an expert panel headed by Splieth found annual restoration failure rates from five to 25 percent at the single tooth level, with high probability of complications in children with multiple teeth treated and overall results ranging from low to unacceptable.^31^

A 2020 systematic review by Schmoeckel summarizes and supports this study’s results with the review findings that “there is high evidence to support a high effect of SDF in arresting, especially cavitated lesions” and “a low level of evidence of moderately high and clinical relevant failure rates for restorative care.^41^”

Given the long wait time and significant occurrence of unplanned visits in the GA group, as well as the negative post-treatment clinical outcomes observed in the SED and GA groups, SDF is a valuable interim treatment alternative. SDF effectively addresses these challenges by providing timely intervention and effective caries management, thereby reducing wait times, and mitigating the risk of negative outcomes. By offering a reliable interim solution, SDF contributes to evidence-based treatment pathways and improves patient experiences while awaiting definitive care and dental rehabilitation.

### Strengths, Limitations, and Generalizability

The strength of this study includes the large and geographically distributed sample size of 4,730 ethnically diverse participants which yielded over 80 percent statistical power. The primary limitation, which negatively impacts the generalizability of this study, is the risk for selection bias with over 90 percent of the participants categorized with Medicaid as the payor source, which serves as a proxy for low socioeconomic status. Hence, the results may not be generalizable to the entire population, since moderate and high socioeconomic status populations were under-represented.

### Future Studies

To reduce selection bias, future studies might broaden socioeconomic sampling breadth by including private practices and dental service organizations as sources of data. Additionally, re-designing future studies as a prospective, longitudinal model would yield long-term clinical outcomes and predictive models of the treated modalities. This would provide valuable insights into the recurrence rate of carious lesions and the potential need for additional intervention, allowing for a better understanding of the optimal timescale for monitoring and managing dental health following various treatment modalities.

## Conclusions

Based upon this study’s results, the following conclusions can be made:

1. Dentists should be aware that:

a. Compared to SDF, the SED and GA groups, respectively, had 1.30- and 2.36-times higher wait-times for treatment completion.
b. Compared to SDF, the SED and GA groups demonstrated higher odds for multiple pre- and post-treatment negative clinical outcomes.
c. With the average age for the respective three groups at 3.43 years (SDF), 4.34 years (SED), and 3.99 years (GA), dentists should adhere to the American Academy of Pediatric Dentistry guideline to encourage the establishment of a dental home for infants by 12 months of age.^42^
c. SDF provides timely and effective caries management with lower wait-time for treatment completion, clinical outcomes consistent with minimally invasive treatment, and mitigation for the risk of negative pre- and post-operative clinical outcomes compared to treatment under sedation or general anesthesia.

## Data Availability

All data produced in the present study are available upon reasonable request to the authors

## Acknowledgments

The authors acknowledge and thank the participants in this study, including the Alaska Native/American Indian peoples who participated through receiving care at the NYU Langone pediatric dentistry training site in Anchorage, Alaska, Hansjorg Wyss Department of Plastic Surgery, NYU Langone Health for funding support and pediatric dental residents Amna Anwar, DDS, Janyn Baird, DDS, Kelsey Coria, DDS, Nikki Crislip, DMD, Joseph Cucolo, DDS, Cory Daley, DMD, Matthew Davidson, DMD, Aleksandra Dragojevic, DDS, Nicole Endo, DDS, Christopher Gordon, DMD, Katherine Gouveia, DMD, Arin Haghverdian, DMD, Nathanael Harrison, DMD, Stephen Hartzog, DMD, Chauncey Hensley, DMD, Louis Jackson, DDS, Melissa Jaramillo, DMD, Yu Jeong Kim, DDS, Alyssa Klaus, DMD, Patricia Larosiliere, DMD, Marc Levicoff, DMD, Horton Li, DMD, Jennifer Macintyre, DDS, Sara Merhi, DDS, Richa Rashmi, DMD, Cody Sia, DMD, James Stewart, DMD, Jacqueline St. Pierre, DMD, MPH, Andrew Stucki, DMD, Cynthia Thomas, DMD, Elizabeth Vaidyan, DMD for data collection contributions.

## Notes

### Competing Interest Statement

The authors have declared no competing interest.

### Funding Statement

This study was funded by the Hansjorg Wyss Department of Plastic Surgery, NYU Langone Health

### Author Declarations

Ethics committee/IRB of the NYU Grossman School of Medicine gave ethical approval for this work

